# Hospital mortality and resource implications of hospitalisation with COVID-19 in London, UK: a prospective cohort study

**DOI:** 10.1101/2020.07.16.20155069

**Authors:** Savvas Vlachos, Adrian Wong, Victoria Metaxa, Sergio Canestrini, Carmen Lopez Soto, Jimstan Periselneris, Kai Lee, Tanya Patrick, Christopher Stovin, Katrina Abernethy, Budoor Albudoor, Rishi Banerjee, Fatima Juma, Sara Al-Hashimi, William Bernal, Ritesh Maharaj

## Abstract

**Background:** Coronavirus disease 2019 (COVID-19) had a significant impact on the National Health Service in the United Kingdom (UK), with over 33 000 cases reported in London by July 6, 2020. Detailed hospital-level information on patient characteristics, outcomes and capacity strain are currently scarce but would guide clinical decision-making and inform prioritisation and planning.

**Methods:** We aimed to determine factors associated with hospital mortality and describe hospital and ICU strain by conducting a prospective cohort study at a tertiary academic centre in London, UK. We included adult patients admitted to hospital with laboratory-confirmed COVID-19 and followed them up until hospital discharge or 30 days. Baseline factors that are associated with hospital mortality were identified via semi-parametric and parametric survival analyses.

**Results:** Our study included 429 patients; 18% of them were admitted to ICU, 52% met criteria for ICU outreach team activation and 61% had treatment limitations placed during their admission. Hospital mortality was 26% and ICU mortality was 34%. Hospital mortality was independently associated with increasing age, male sex, history of chronic kidney disease, increasing baseline C-reactive protein level and dyspnoea at presentation. COVID-19 resulted in substantial ICU and hospital strain, with up to 9 daily ICU admissions and 41 daily hospital admissions, to a peak census of 80 infected patients admitted in ICU and 250 in the hospital. Management of such a surge required extensive reorganisation of critical care services with expansion of ICU capacity from 69 to 129 beds, redeployment of staff from other hospital areas and coordinated hospital-level effort.

**Conclusions:** COVID-19 is associated with a high burden of mortality for patients treated on the ward and the ICU and required substantial reconfiguration of critical care services. This has significant implications for planning and resource utilization.

## INTRODUCTION

Coronavirus Disease 2019 (COVID-19), an infectious syndrome caused by SARS-CoV-2, appeared in December 2019 and evolved into a pandemic that caused more than eleven million cases and 530 000 deaths worldwide by July 2020 [1]. The high number and acuity of patients resulted in unprecedented demand for hospitalisation and critical care services in many affected countries. Developed areas such as Wuhan (China), Lombardy (Italy), and New York (United States; USA) reported a surge in critically ill patients, which quickly led to significant strain on healthcare systems through shortages in Intensive Care Unit (ICU) beds, equipment, and trained personnel [2-4].

There was concern that the United Kingdom (UK) would face similar challenges, particularly in densely populated areas like London. The first infection was reported on January 30, 2020 and by early July the country had recorded a large number of cases, with over 33 000 confirmed infections in the greater area of London alone [5].

Despite the high number of hospital admissions with COVID-19, the existing peer-reviewed literature in the UK remains restricted to large population-level studies and small retrospective cohorts with few details on clinical management and hospital-level strain [6-8]. Major reports focus exclusively either on the pre-ICU stage of illness [7] or the ICU management [8] and follow-up times are generally short [7]. Extrapolation from international settings is difficult due significant differences in population characteristics [2], health system organisation [2,4,9] and health system strain [10].

Detailed local patient-level information on characteristics and outcomes as well as institution-level information on service pressures would guide clinical decision making and inform effective prioritisation and resource allocation in the future, particularly in circumstances when a surge in demand places the National Health Service again at risk of being overwhelmed.

The aim of our study was to describe the clinical characteristics and course of hospitalised patients with COVID-19, as well as its broader resource implications for a tertiary academic hospital in London, UK.

## METHODS

### Study design

We conducted a prospective cohort study at King’s College Hospital (KCH), a tertiary academic centre in London, UK. Institutional (IRAS-256619; April 15, 2020) and regional (Health Research Authority; IRAS-256619; April 22, 2020) review board approvals waived the need for ethics committee review and the need for informed patient consent.

All consecutive patients tested for SARS-CoV-2 infection using reverse-transcriptase polymerase chain reaction (RT-PCR) assays of respiratory tract samples between February 25 and March 31, 2020 were considered eligible for study participation. Inclusion criteria were age of 18 years or above and laboratory-confirmed SARS-CoV-2 infection, which was defined as at least one positive RT-PCR nasopharyngeal swab [11]. Patients with missing identifiers, missing SARS-CoV-2 test results, and those transferred from other hospitals were excluded. Included patients were followed up until death, hospital discharge, or 30 days after hospital admission. Follow-up was concluded on April 30, 2020. For patients with multiple hospital or ICU admissions, only the first admission was recorded. Transfers between different ICUs within the hospital were considered part of the same admission. The primary outcome was mortality at hospital discharge or 30 days. Secondary outcomes were ICU mortality, as well as hospital and ICU capacity strain, measured in bed occupancy. The report of our findings is based on the Strengthening the Reporting of Observational Studies in Epidemiology (STROBE) Statement [12]. More details regarding the institutional setting and pandemic surge are available in the Supplementary file (page 1).

### Data collection and management

Trained members of the clinical team extracted anonymised data from electronic health records (EHR) [13]. We recorded the following for all included patients: age, sex, ethnicity, area-level socioeconomic deprivation (Index of Multiple Deprivation [14]) clinical frailty, medical comorbidities, age-adjusted Charlson Comorbidity Index (ACCI) body mass index (BMI), prior residence, self-reported presenting symptoms, reason for hospital admission, laboratory, microbiological and imaging tests, COVID-19-specific drug treatments, treatment limitations such as Treatment Escalation Plans (TEPs) or Do-Not-Resuscitate (DNR) orders, length of hospital stay and outcome at hospital discharge or at 30 days.

For patients who were initially admitted to the ward we also recorded peak temperature and National Early Warning Score version 2 (NEWS2) score [15], maximum level of respiratory support and development of complications such as hypoxia, hypotension, tachycardia, and depressed consciousness. For patients who were admitted to ICU we additionally recorded reason for ICU admission, daily Sequential Organ Failure Assessment (SOFA) score [16], organ support (mechanical ventilation, renal replacement therapy, circulatory support), arterial partial pressure of oxygen (P_a_O_2_), fraction of inspired oxygen (F_i_O_2_), P_a_O_2_/F_i_O_2_ ratio (PFR), use of medications such as vasopressors, pulmonary vasodilators and neuromuscular blocking drugs, use of prone positioning or Extracorporeal Membrane Oxygenation (ECMO), insertion of tracheostomy, length of ICU stay, and outcome at ICU discharge or at 30 days.

We recorded comorbidities and symptoms based on electronic case note review and categorised variables according to clinical relevance and current literature [4,10,17]. We used only validated laboratory results and official reports of imaging studies. Initial tests for newly admitted COVID-19 patients refer to those performed within 24 hours of hospital admission; for patients already admitted, they refer to tests performed within 24 hours of COVID-19 diagnosis. Clinical frailty was assessed on a 9-category scale [18], using information available in the EHR; patients with a score above 4 were considered frail [19]. Treatment escalation plans (TEPs) refer to structured assessments of patients’ suitability regarding specific aspects of treatment such as organ support or ICU admission [20]. Acute Kidney Injury (AKI) was defined according to Kidney-Disease Improving Global Outcomes (KDIGO) criteria [21]. More details regarding definitions of collected data are provided in the Supplementary file (pages 2-3).

### Statistical methods

Descriptive analyses are presented as median (IQR [range]) or number (%) and we avoided univariate comparisons between groups. To identify factors associated with the time to death at hospital discharge or at 30 days, we performed multivariable Cox proportional-hazards and parametric survival analyses. Covariate inclusion followed a structured approach. We tested the proportional hazards assumption and investigated interactions of the included covariates with sex and age. We also assessed whether the baseline survival experience differed by categories of age, sex, frailty and ethnicity, using stratified Cox regression. The effect of each included covariate was quantified by calculating adjusted hazard ratios (HRs) with 95% Confidence Intervals (95%CI). We followed a similar approach for the parametric analysis and plotted the hazard function over time for six hypothetical patients, in order to show the effect of each included covariate on the hazard of death. Statistical tests were 2-sided, with an α-level of 0.05 for statistical significance. We did not impute any missing data. Analyses were performed using Stata/MP version 15.1 (StataCorp). Details regarding the statistical approach are provided in the Supplementary file (pages 4-10).

## RESULTS

Between February 25th and March 31st, 2020, 2728 patients were tested for SARS-CoV-2 infection at KCH. After excluding 170 patients (6%), 2558 patients (94%) were screened for inclusion; 2129 (83%) of them did not meet the inclusion criteria and 429 (17%) were included in the study. Among them, 353 patients (82%) were treated only on the ward and 76 (18%) were treated in ICU. The study flow diagram is shown in Figure 1.

**Figure 1:**
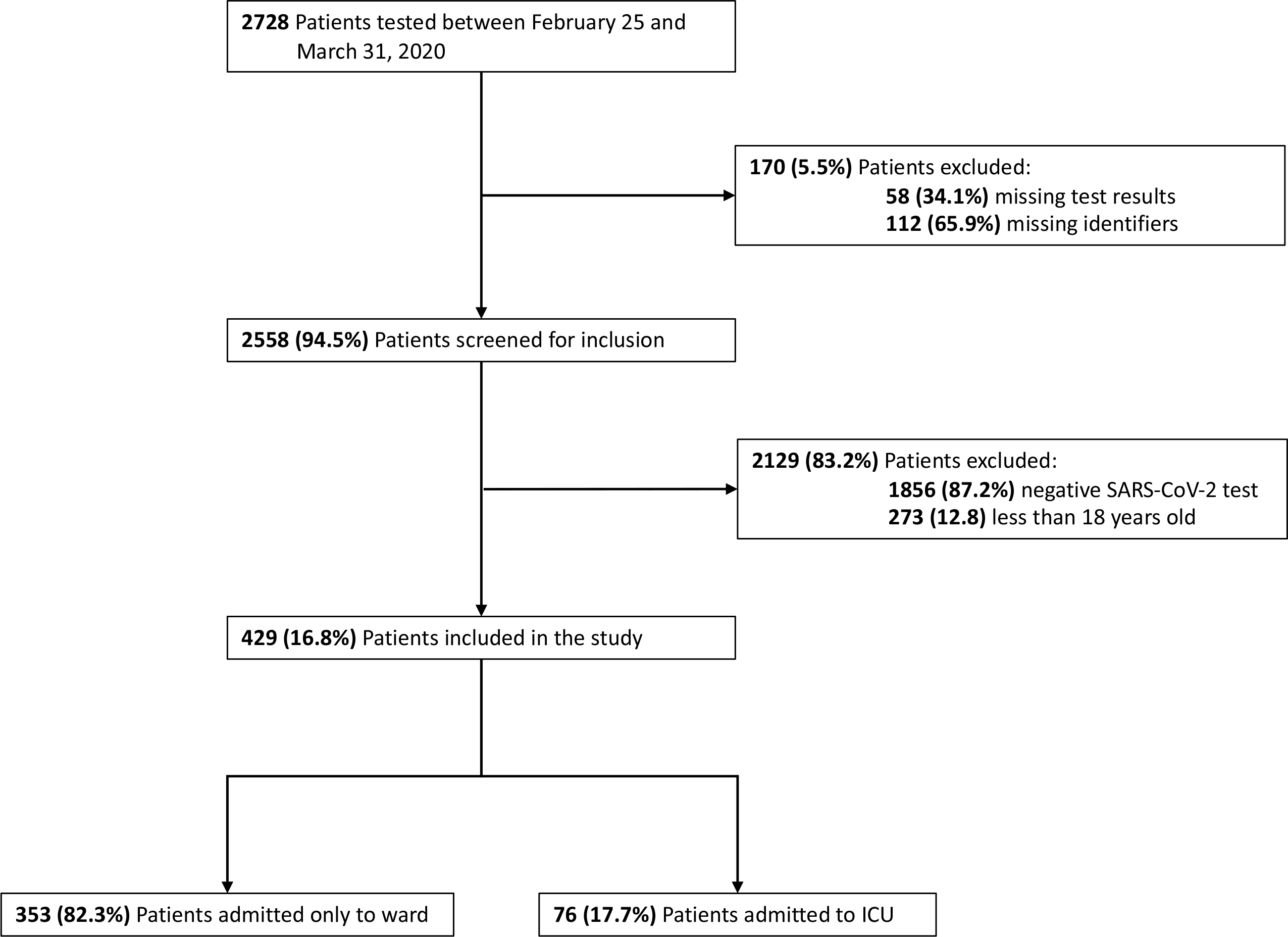
The study flow diagram.

Baseline patient characteristics are shown in Table 1. Patients had a median age of 65 years (IQR 52-81 [18-97]) and 61% were of ethnic minority background. The majority (61%) were overweight and 90% of them had at least one serious comorbidity; the most common were hypertension (52%), diabetes (37%), and chronic kidney disease (CKD) (14%). Most common presenting complaints included cough (62%), fever (62%), and dyspnoea (43%), but only 42% of patients were hospitalised for hypoxaemic respiratory failure. Initial chest x-rays were normal in 23% of patients and showed diffuse, bilateral infiltrates in more than half. On admission, lymphopenia was common (Table 2), as were elevations in C-reactive protein (CRP) (median 75 mg.l^-1^, IQR 28-143 [0-608]) and creatinine (median 92 mg.l^-1^, IQR 68-130 [25-1000]). Raised Lactate Dehydrogenase, Ferritin, Creatine Kinase and D-dimers were also common, albeit measured in a minority of patients. Compared to ward patients, those admitted to ICU were younger, less commonly frail and had a lower burden of chronic comorbidities, as described by the ACCI. They were, however, more likely to be diabetic and to be hospitalised for respiratory failure, with significantly more deranged initial laboratory tests and abnormal chest imaging.

**Table 1:** Baseline characteristics of patients, by admission location. Data are reported as median (IQR [range]) or n (%), or n/N (%) when some data are missing.

|  | Study population<br>(n=429) | Treated only on ward<br>(n=353) | Treated in ICU<br>(n=76) |
| --- | --- | --- | --- |
| Age; years | 65 (52-81 [18-97]) | 68 (54-82 [18-97]) | 57 (48-63 [25-79]) |
| 18-29 | 12 (3%) | 9 (2%) | 3 (4%) |
| 30-39 | 27 (6%) | 21 (6%) | 6 (8%) |
| 40-49 | 37 (9%) | 26 (7%) | 11 (14%) |
| 50-59 | 79 (18%) | 56 (16%) | 23 (30%) |
| 60-69 | 90 (21%) | 69 (19%) | 21 (28%) |
| 70-79 | 64 (15%) | 55 (16%) | 9 (12%) |
| 80-89 | 82 (19%) | 79 (22%) | 3 (4%) |
| ≥ 90 | 38 (9%) | 38 (11%) | 0 (0%) |
| Sex |  |  |  |
| Female | 195 (45%) | 169 (48%) | 26 (34%) |
| Male | 234 (55%) | 184 (52%) | 50 (66%) |
| Ethnicity * |  |  |  |
| Black | 187/391 (48%) | 148 (47%) | 39 (53%) |
| Asian | 15/391 (4%) | 9 (3%) | 6 (8%) |
| White | 151/391 (38%) | 129 (40%) | 22 (30%) |
| Mixed or other | 38/391 (10%) | 31 (10%) | 7 (9%) |
| BMI † | 26.6 (23.2-31.2 [14-94]) | 26.2 (22.8-31.0 [14-49]) | 27.8 (24.2-32.0 [20-40]) |
| ≤ 25 | 115/296 (39%) | 90/222 (40%) | 25/74 (34%) |
| 26-30 | 91/296 (31%) | 69/222 (31%) | 22/74 (30%) |
| 21-35 | 54/296 (18%) | 36/222 (16%) | 18/74 (24%) |
| 36-40 | 18/296 (6%) | 13/222 (6%) | 5/74 (7%) |
| ≥ 40 | 18/296 (6%) | 14/222 (6%) | 4/74 (5%) |
| ACCI ‡ | 4 (2-6 [0-15]) | 4 (2-6 [0-15]) | 2 (1-4 [0-12]) |
| 0 | 40/401 (10%) | 33/33 (10%) | 7/70 (10%) |
| 1 | 49/401 (12%) | 34 (10%) | 15 (21%) |
| 2 | 51/401 (13%) | 37 (11%) | 14 (20%) |
| ≥3 | 261/401 (65%) | 227 (69%) | 34 (49%) |
| IMD quintile § |  |  |  |
| 1 (Least deprived) | 9/427 (2%) | 6/351 (2%) | 3 (4%) |
| 2 | 36/427 (8%) | 31/351 (9%) | 5 (7%) |
| 3 | 86/427 (20%) | 72/351 (20%) | 14 (18%) |
| 4 | 204 /427 (48%) | 167/351 (48%) | 37 (49%) |
| 5 (Most deprived) | 92/427 (21%) | 75/351 (21%) | 17 (22%) |
| Prior residence |  |  |  |
| Home | 382 (89%) | 308 (87%) | 74 (97%) |
| Nursing home | 31 (7%) | 30 (8%) | 1 (1.5%) |
| Health-related institution | 6 (1%) | 6 (2%) | 0 (0%) |
| Other | 10 (2%) | 9 (3%) | 1 (1.5%) |
| Comorbidities |  |  |  |
| Hypertension | 225 (52%) | 187 (53%) | 38 (50%) |
| Diabetes mellitus | 161 (37%) | 122 (35%) | 39 (51%) |
| Coronary heart disease | 41 (10%) | 36 (10%) | 5 (7%) |
| Chronic heart failure | 28 (6%) | 28 (8%) | 0 (0%) |
| Chronic kidney disease | 61 (14%) | 54 (15%) | 7 (9%) |
| End-stage renal disease | 14 (3%) | 12 (3%) | 2 (3%) |
| Chronic respiratory disease | 51 (12%) | 43 (12%) | 8 (10%) |
| Chronic liver disease | 8 (2%) | 6 (2%) | 2 (3%) |
| Cerebrovascular accident | 57 (13%) | 55 (16%) | 2 (3%) |
| Immunosuppression (incl. HIV) | 26 (6%) | 25 (7%) | 1 (1%) |
| Sickle cell disease | 7 (2%) | 5 (1%) | 2 (3%) |
| Active cancer or haematological disease | 41 (10%) | 39 (11%) | 2 (3%) |
| Frailty |  |  |  |
| Frail | 170 (40%) | 156/351 (44%) | 14 (18%) |
| Not frail | 259 (60%) | 197/351 (56%) | 62 (82%) |
| Chief presenting symptoms |  |  |  |
| Fever | 265 (62%) | 212 (60%) | 53 (70%) |
| Cough | 266 (62%) | 212 (60%) | 54 (71%) |
| Dyspnoea | 184 (43%) | 143 (40%) | 41 (54%) |
| Fatigue | 101 (23%) | 80 (23%) | 21 (28%) |
| Lethargy | 82 (19%) | 67 (19%) | 15 (20%) |
| Days since symptom onset ¶ | 3.5 (1-6 [0-37]) | 3 (1-6 [0-37]) | 4.5 (2-6 [0-14]) |
| Reason for hospital admission |  |  |  |
| Medical (hypoxemic respiratory failure) | 182 (42%) | 125 (35%) | 57 (75%) |
| Medical (other reason) | 215 (50%) | 203 (57%) | 12 (16%) |
| Surgical or trauma | 29 (7%) | 22 (6%) | 7 (9%) |
| Elective | 3 (1%) | 3 (1%) | 0 (0%) |
| Initial chest x-ray |  |  |  |
| No abnormal findings | 95/405 (23%) | 88/329 (27%) | 7 (9%) |
| Diffuse opacities | 155/310 (50%) | 105/241 (44%) | 50/69 (72%) |
| Bilateral opacities | 178/310 (57%) | 124/241 (51%) | 54/69 (78%) |
| Pleural effusion | 42/310 (13%) | 33/241 (14%) | 9/69 (13%) |

Abbreviations: ICU, Intensive Care Unit; BMI, Body Mass Index (calculated as weight in kilograms divided by height in meters squared); ACCI, Age-adjusted Charlson Comorbidity Index; IMD, Index of Multiple Deprivation; HIV, Human Immunodeficiency Virus.
\* Ethnicity was missing in 38 patients (9%)
† BMI was missing in 133 patients (31%)
‡ ACCI was missing for 28 patients (6%)
§ IMD was missing in 2 patients (<1%)
¶ Data on symptom onset was missing in 217/429 (51%) of patients

**Table 2:**
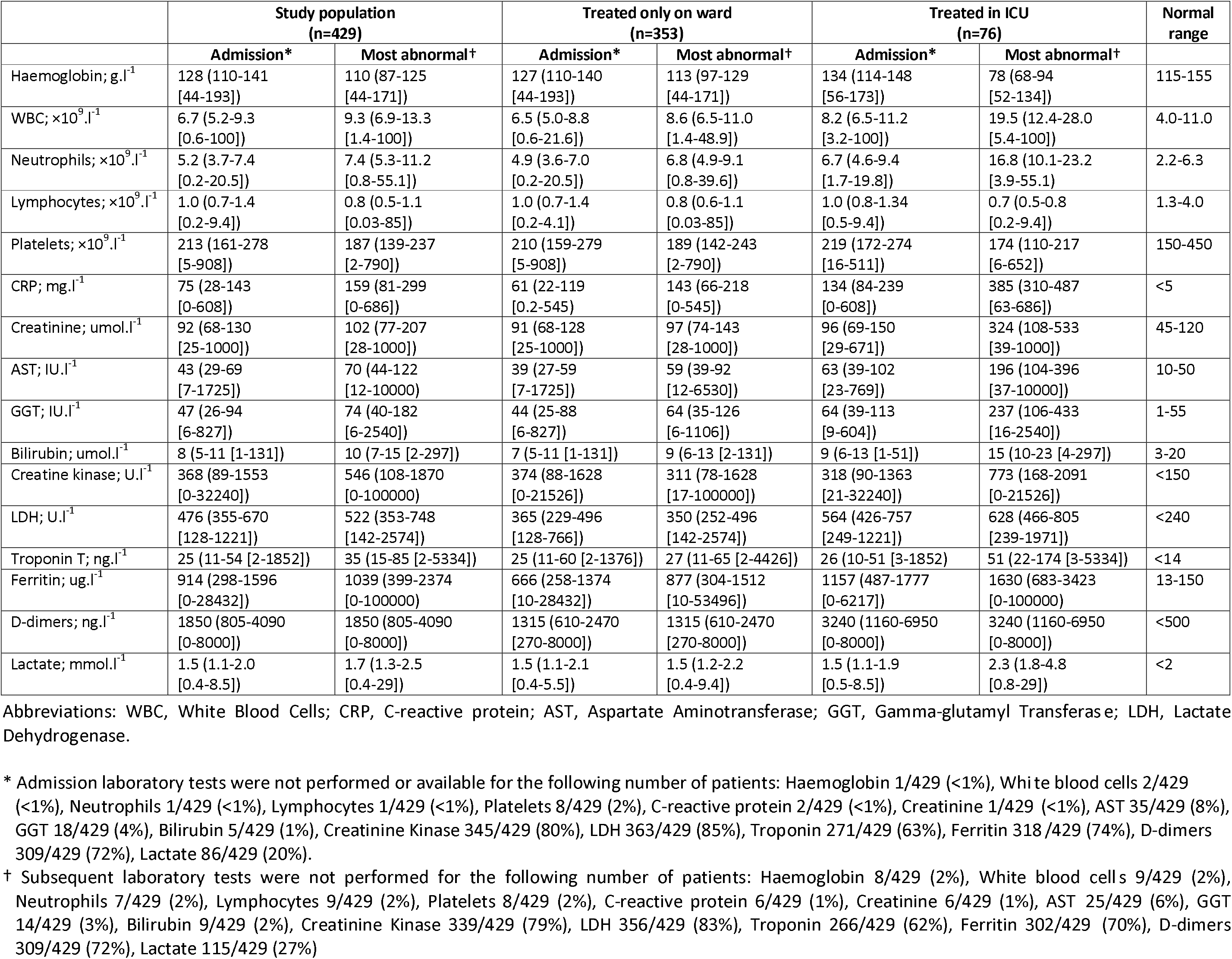
Admission and most abnormal laboratory values for the study population, by location of admission. Data are reported as median (IQR [range])

### Patients treated on the ward

Most patients developed significant morbidity during their hospital admission. Fifty-two percent met criteria for critical care outreach team activation (NEWS2 score above six), predominantly for respiratory failure (Table 3). On chest computed tomography (CT), more than 70% of scans showed bilateral diffuse infiltrates and 25% revealed pulmonary embolism (PE). PE was more frequently identified in ICU patients (33%). Laboratory abnormalities (Table 2) included worsening lymphopenia, raised CRP (median 159 mg.l^-1^, IQR 81-299 [0-686]) and intracellular enzymes, as well as impaired hepatic, renal, and haemostatic function. Overall, 21% of patients developed KDIGO stage three AKI but the incidence among ICU patients (67%) was much higher than that among ward patients (11%). We recorded only mild degrees of myocardial involvement, evidenced by small increases in values of high-sensitivity Troponin T and unremarkable echocardiographic findings. Secondary infections were common in the ICU-treated group: 50% had a positive respiratory tract sample and 41% had a positive blood culture. Samples from the respiratory tract were frequently positive for Gram negative organisms and fungi.

**Table 3:**
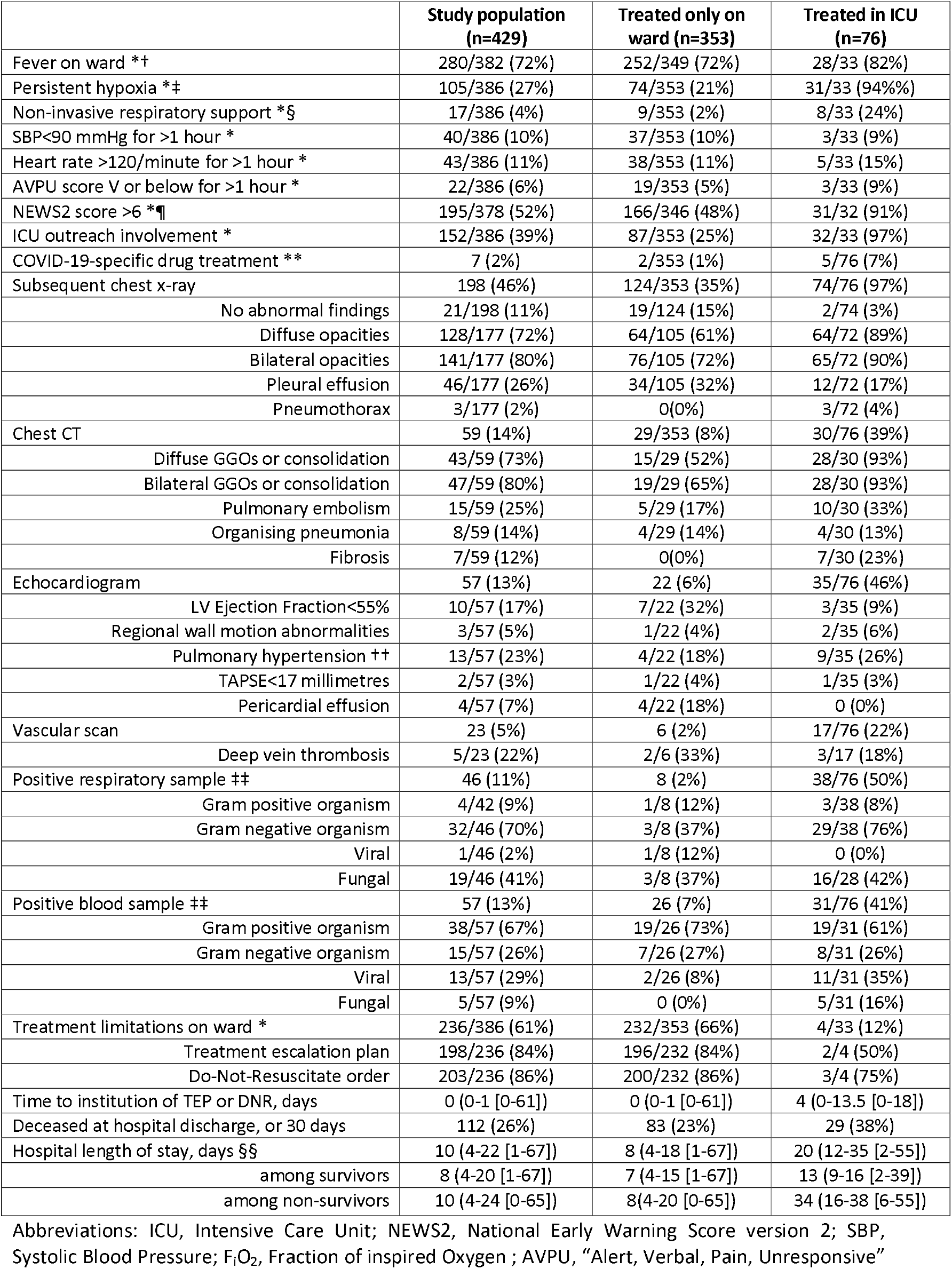
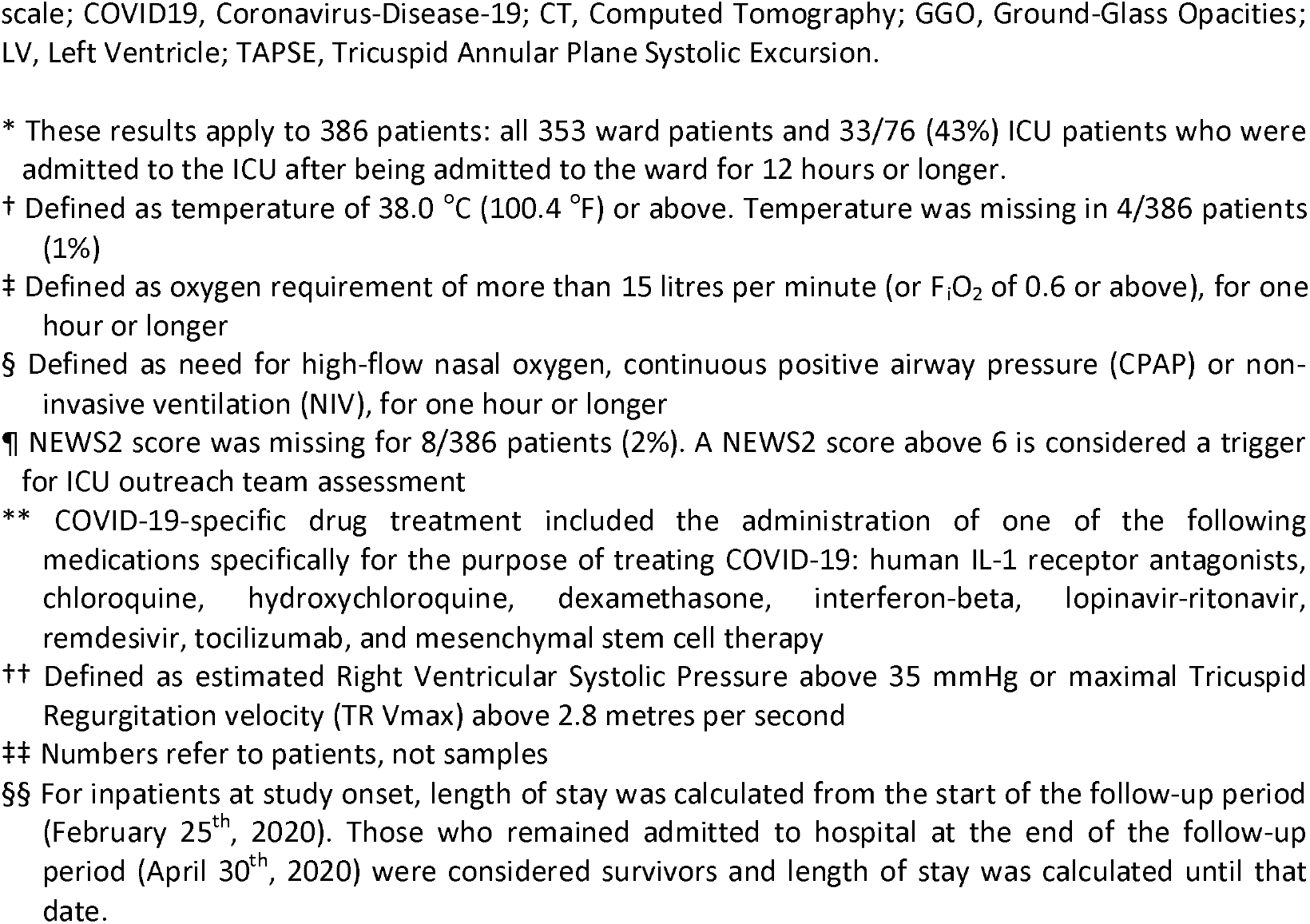
Clinical findings and outcomes during the hospital admission. Data are reported as median (IQR [range]) or n (%), or n/N (%) when some data are missing.

|  | Study population<br>(n=429) | Treated only on<br>ward (n=353) | Treated in ICU<br>(n=76) |
| --- | --- | --- | --- |
| Fever on ward *† | 280/382 (72%) | 252/349 (72%) | 28/33 (82%) |
| Persistent hypoxia *‡ | 105/386 (27%) | 74/353 (21%) | 31/33 (94%) |
| Non-invasive respiratory support *§ | 17/386 (4%) | 9/353 (2%) | 8/33 (24%) |
| SBP<90 mmHg for >1 hour * | 40/386 (10%) | 37/353 (10%) | 3/33 (9%) |
| Heart rate >120/minute for >1 hour * | 43/386 (11%) | 38/353 (11%) | 5/33 (15%) |
| AVPU score V or below for >1 hour * | 22/386 (6%) | 19/353 (5%) | 3/33 (9%) |
| NEWS2 score >6 *¶ | 195/378 (52%) | 166/346 (48%) | 31/32 (91%) |
| ICU outreach involvement * | 152/386 (39%) | 87/353 (25%) | 32/33 (97%) |
| COVID-19-specific drug treatment ** | 7 (2%) | 2/353 (1%) | 5/76 (7%) |
| Subsequent chest x-ray | 198 (46%) | 124/353 (35%) | 74/76 (97%) |
| No abnormal findings | 21/198 (11%) | 19/124 (15%) | 2/74 (3%) |
| Diffuse opacities | 128/177 (72%) | 64/105 (61%) | 64/72 (89%) |
| Bilateral opacities | 141/177 (80%) | 76/105 (72%) | 65/72 (90%) |
| Pleural effusion | 46/177 (26%) | 34/105 (32%) | 12/72 (17%) |
| Pneumothorax | 3/177 (2%) | 0(0%) | 3/72 (4%) |
| Chest CT | 59 (14%) | 29/353 (8%) | 30/76 (39%) |
| Diffuse GGOs or consolidation | 43/59 (73%) | 15/29 (52%) | 28/30 (93%) |
| Bilateral GGOs or consolidation | 47/59 (80%) | 19/29 (65%) | 28/30 (93%) |
| Pulmonary embolism | 15/59 (25%) | 5/29 (17%) | 10/30 (33%) |
| Organising pneumonia | 8/59 (14%) | 4/29 (14%) | 4/30 (13%) |
| Fibrosis | 7/59 (12%) | 0(0%) | 7/30 (23%) |
| Echocardiogram | 57 (13%) | 22 (6%) | 35/76 (46%) |
| LV Ejection Fraction<55% | 10/57 (17%) | 7/22 (32%) | 3/35 (9%) |
| Regional wall motion abnormalities | 3/57 (5%) | 1/22 (4%) | 2/35 (6%) |
| Pulmonary hypertension †† | 13/57 (23%) | 4/22 (18%) | 9/35 (26%) |
| TAPSE<17 millimetres | 2/57 (3%) | 1/22 (4%) | 1/35 (3%) |
| Pericardial effusion | 4/57 (7%) | 4/22 (18%) | 0 (0%) |
| Vascular scan | 23 (5%) | 6 (2%) | 17/76 (22%) |
| Deep vein thrombosis | 5/23 (22%) | 2/6 (33%) | 3/17 (18%) |
| Positive respiratory sample ‡‡ | 46 (11%) | 8 (2%) | 38/76 (50%) |
| Gram positive organism | 4/42 (9%) | 1/8 (12%) | 3/38 (8%) |
| Gram negative organism | 32/46 (70%) | 3/8 (37%) | 29/38 (76%) |
| Viral | 1/46 (2%) | 1/8 (12%) | 0 (0%) |
| Fungal | 19/46 (41%) | 3/8 (37%) | 16/28 (42%) |
| Positive blood sample ‡‡ | 57 (13%) | 26 (7%) | 31/76 (41%) |
| Gram positive organism | 38/57 (67%) | 19/26 (73%) | 19/31 (61%) |
| Gram negative organism | 15/57 (26%) | 7/26 (27%) | 8/31 (26%) |
| Viral | 13/57 (29%) | 2/26 (8%) | 11/31 (35%) |
| Fungal | 5/57 (9%) | 0 (0%) | 5/31 (16%) |
| Treatment limitations on ward * | 236/386 (61%) | 232/353 (66%) | 4/33 (12%) |
| Treatment escalation plan | 198/236 (84%) | 196/232 (84%) | 2/4 (50%) |
| Do-Not-Resuscitate order | 203/236 (86%) | 200/232 (86%) | 3/4 (75%) |
| Time to institution of TEP or DNR, days | 0 (0-1 [0-61]) | 0 (0-1 [0-61]) | 4 (0-13.5 [0-18]) |
| Deceased at hospital discharge, or 30 days | 112 (26%) | 83 (23%) | 29 (38%) |
| Hospital length of stay, days §§ | 10 (4-22 [1-67]) | 8 (4-18 [1-67]) | 20 (12-35 [2-55]) |
| among survivors | 8 (4-20 [1-67]) | 7 (4-15 [1-67]) | 13 (9-16 [2-39]) |
| among non-survivors | 10 (4-24 [0-65]) | 8(4-20 [0-65]) | 34 (16-38 [6-55]) |

Abbreviations: ICU, Intensive Care Unit; NEWS2, National Early Warning Score version 2; SBP, Systolic Blood Pressure; F<sub>I</sub>O<sub>2</sub>, Fraction of inspired Oxygen ; AVPU, “Alert, Verbal, Pain, Unresponsive”
scale; COVID19, Coronavirus-Disease-19; CT, Computed Tomography; GGO, Ground-Glass Opacities; LV, Left Ventricle; TAPSE, Tricuspid Annular Plane Systolic Excursion.
\* These results apply to 386 patients: all 353 ward patients and 33/76 (43%) ICU patients who were admitted to the ICU after being admitted to the ward for 12 hours or longer.
† Defined as temperature of 38.0 °C (100.4 °F) or above. Temperature was missing in 4/386 patients (1%)
‡ Defined as oxygen requirement of more than 15 litres per minute (or $F_{iO_2}$ of 0.6 or above), for one hour or longer
§ Defined as need for high-flow nasal oxygen, continuous positive airway pressure (CPAP) or non-invasive ventilation (NIV), for one hour or longer
¶ NEWS2 score was missing for 8/386 patients (2%). A NEWS2 score above 6 is considered a trigger for ICU outreach team assessment
\*\* COVID-19-specific drug treatment included the administration of one of the following medications specifically for the purpose of treating COVID-19: human IL-1 receptor antagonists, chloroquine, hydroxychloroquine, dexamethasone, interferon-beta, lopinavir-ritonavir, remdesivir, tocilizumab, and mesenchymal stem cell therapy
†† Defined as estimated Right Ventricular Systolic Pressure above 35 mmHg or maximal Tricuspid Regurgitation velocity (TR Vmax) above 2.8 metres per second
‡‡ Numbers refer to patients, not samples

Treatment limitations were placed in 61% of patients overall, most commonly on hospital admission (median 0 days, IQR 0-1 [0-61]). The majority (>80%) of these limitations involved TEPs and DNR orders. Two-thirds of patients with a TEP were considered frail. Among patients with treatment limitations that were not considered frail, many had significant medical comorbidities such as active malignancy and stroke.

### Patients treated in ICU

The clinical trajectory of 76 patients who were admitted to ICU is described in Table 4. Many patients developed precipitous respiratory failure and required ICU admission directly from the emergency department (57%). Multiple organ failure was common with 97% requiring mechanical ventilation for hypoxic respiratory failure, 80% requiring pharmacologic circulatory support and 57% requiring renal replacement therapy (RRT) during their ICU stay. The severity of illness was reflected in the high SOFA scores, which remained elevated even after two weeks of ICU stay. The median duration of mechanical ventilation was 12 days (IQR 6-23 [1-37]) and that of renal replacement was 11 days (IQR 4-17 [1-37]). Patients frequently required rescue oxygenation strategies (54%), which included neuromuscular blocking drugs, inhaled prostacyclin and prone positioning. Extracorporeal membrane oxygenation (ECMO) was used only in 5% of ICU patients. A significant proportion (38%) of patients required a tracheostomy, which was performed at a median of 16.5 days (IQR 14-20 [10-38]) after ICU admission.

**Table 4:** Patient management and outcomes in the Intensive Care Unit. Data are reported as median (IQR [range]) or n (%)

|  | Study population (n=76) |
| --- | --- |
| Fever in the ICU * | 57 (78%) |
| CPAP or NIV | 10 (13%) |
| Mechanical ventilation | 74 (97%) |
| Duration; days | 12 (6-23 [1-37]) |
| Renal replacement therapy | 43 (57%) |
| Duration; days | 11 (4-17 [1-37]) |
| Pharmacological circulatory support † | 61 (80%) |
| Duration; days | 8 (4-14 [1-39]) |
| SOFA score ‡ |  |
| Day 1 (admission) | 14 (13-16 [7-20]) |
| Day 3 | 15 (12-17 [7-20]) |
| Day 7 | 15 (13-17 [9-20]) |
| Day 10 | 15 (14-16 [8 [7-20]-20]) |
| Day 14 | 14 (12-17 [8-20]) |
| Highest F<sub>I</sub>O<sub>2</sub> ‡ |  |
| Day 1 (admission) | 0.6 (0.5-0.8 [0.3-1]) |
| Day 3 | 0.5 (0.4-0.6 [0.28-1]) |
| Day 7 | 0.5 (0.4-0.7 [0.25-1]) |
| Day 10 | 0.5 (0.4-0.7 [0.21-1]) |
| Day 14 | 0.4 (0.3-0.7 [0.21-1]) |
| Lowest P<sub>a</sub>O<sub>2</sub>/F<sub>I</sub>O<sub>2</sub> ratio; kPa ‡ |  |
| Day 1 (admission) | 17.5 (13.7-25 [8-60]) |
| Day 3 | 19.2 (14-24.7 [8-42.5]) |
| Day 7 | 19 (12.5-27.5 [7-45]) |
| Day 10 | 20.5 (15-29 [7-46]) |
| Day 14 | 23.7 (14-30.2 [9-53]) |
| Rescue oxygenation strategies | 41 (54%) |
| Neuromuscular blocking drugs | 31 (41%) |
| Inhaled prostacyclin | 22 (29%) |
| Prone positioning | 19 (25%) |
| ECMO | 4 (5%) |
| Tracheostomy | 27 (38%) |
| Time to tracheostomy; days | 16.5 (14-20 [10-38]) |
| Treatment limitations in ICU | 18 (24%) |
| Do-Not-Resuscitate (DNR) order | 15 (20%) |
| Time to DNR order; days | 6.5 (4-13 [0-22]) |
| Treatment escalation plan (TEP) | 16 (21%) |
| Time to TEP order; days | 7.5 (3.5-12.5 [0-36]) |
| Treatment withdrawal | 8 (10%) |
| Time to withdrawal; days | 12 (4.5-18 [3-36]) |
| Deceased at ICU discharge, or 30 days | 26 (34%) |
| ICU length of stay; days § | 13 (7-30 [1-51]) |
| among survivors | 19 (9-32 [2-51]) |
| among non-survivors | 11 (5-14 [1-39]) |

Abbreviations: CPAP: Continuous Positive Airway Pressure; NIV: Non-invasive Ventilation; SOFA: Sequential Organ Failure Assessment; PaO<sub>2</sub>: Partial pressure of oxygen in arterial blood; F<sub>I</sub>O<sub>2</sub>: Fraction of Inspired Oxygen; ECMO: Extracorporeal Membrane Oxygenation; ICU: Intensive Care Unit.
\* Defined as temperature of 38.3 °C (101 °F) or above
† Defined as use of any vasopressor or inotropic drug
‡ Data are missing for 1 patient (1%)
§ Those who remained admitted to hospital at the end of the follow-up period (April 30th, 2020) were considered survivors and length of stay was calculated until that date.

### Outcomes

Unadjusted mortality at hospital discharge or 30 days was 26% overall, 23% for ward patients and 38% for those treated in ICU. In unadjusted Cox survival analysis, hospital mortality was associated age, sex, ACCI score, frailty category, CRP, creatinine, CKD, diabetes, and dyspnoea or fever as presenting complaints. After adjustment, it was independently associated with increasing age (HR 1.07 per decade above 40 years; 95%CI 1.04-1.09, p<0.001); male sex (HR 2.31; 95%CI 1.52-3.50, p<0.001); raised admission CRP level (HR 1.03 per 10 mg.l^-1^ increments above the upper normal limit of 5 mg.l^-1^; 95%CI 1.01-1.04, p=0.001); history of CKD (HR 1.87; 95%CI 1.21-2.89, p=0.005) and dyspnoea as a presenting symptom (HR 1.88; 95%CI 1.24-1.86, p=0.003). Ethnicity or level of deprivation were not associated with mortality in unadjusted or adjusted analyses. Stratification of the model by categories of age (≤60, >60 years), sex, frailty or ethnicity did not provide evidence of differing baseline hazard. The effect of each included covariate, based on parametric modelling, is shown in Figure 2 with six examples of hypothetical patients. In the parametric model, male sex and dyspnoea on presentation had the largest impact on the hazard function. More details regarding the results of the parametric analysis are available in the Supplementary file (pages 8-10).

**Figure 2:**
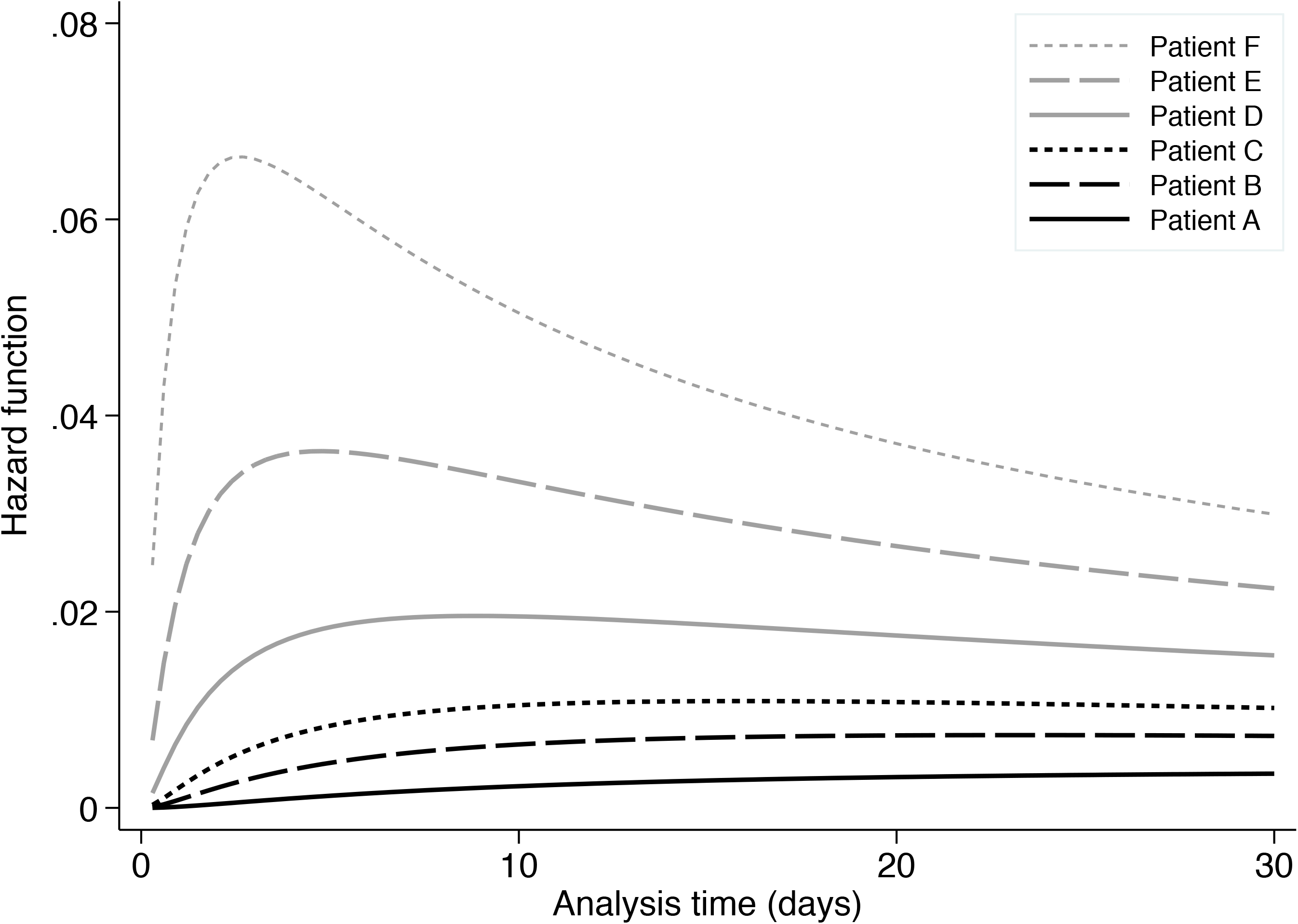
A plot of the primary outcome (hazard of death at hospital discharge or 30 days) over time for six hypothetical patients, based on the parametric survival analysis. Patients characteristics were adjusted to highlight the additional effect of individual risk factors on the hazard of death. Patient A represents a female 40-year old patient with normal C-reactive protein levels (CRP <5 mg.l^-1^), no Chronic Kidney Disease (CKD) and no dyspnoea on presentation. Patient B represents a male 40-year old patient with normal CRP, no CKD and no dyspnoea. Patient C represents a 65-year old male patient with normal CRP, no CKD and no dyspnoea. Patient D represents a 65-year old male patient with an abnormal CRP of 200 mg.l^-1^, no CKD and no dyspnoea. Patient E is similar with Patient D but has a history of CKD. Finally, Patient F represents a 65-year old male patient with an abnormal CRP of 200 mg.l^-1^, a history of CKD and presentation to hospital with dyspnoea.

At the end of the 30-day follow-up, 15 of 353 ward patients (4%) remained admitted in hospital and 12 of 270 (4%) were readmitted after hospital discharge. Among all ICU patients, 13 (17%) remained admitted in ICU at the end of follow-up. Among the 50 patients discharged from ICU, three (6%) were readmitted to ICU, 25 (50%) were discharged from hospital, three (6%) died on the ward and 22 (44%) remained hospitalised on the ward.

During the study period, the hospital experienced a sudden surge in critically ill patients and details regarding the number of hospital and ICU admissions are shown in Figure 3. Over a period of approximately six weeks, 20-40 patients were admitted daily to the hospital with COVID-19 and between five to ten of them required admission to ICU for organ support. At the peak of the pandemic, both the hospital and ICUs experienced significant capacity strain, with 28% of the hospital capacity of 900 beds and 70% of the ICU surge capacity of 129 beds taken up by patients with COVID-19. This increased demand was supported by changes to the service described in Table 5. Major changes included Anaesthetic cover for newly opened ICU beds, re-deployment of the entire hospital workforce to support ICU, delivery of critical care in non-conventional areas like operating rooms, and the introduction of teams dedicated to tasks like prone positioning, communication with families, and tracheostomies. More details are available in the Supplementary file (page 11).

**Table 5:**
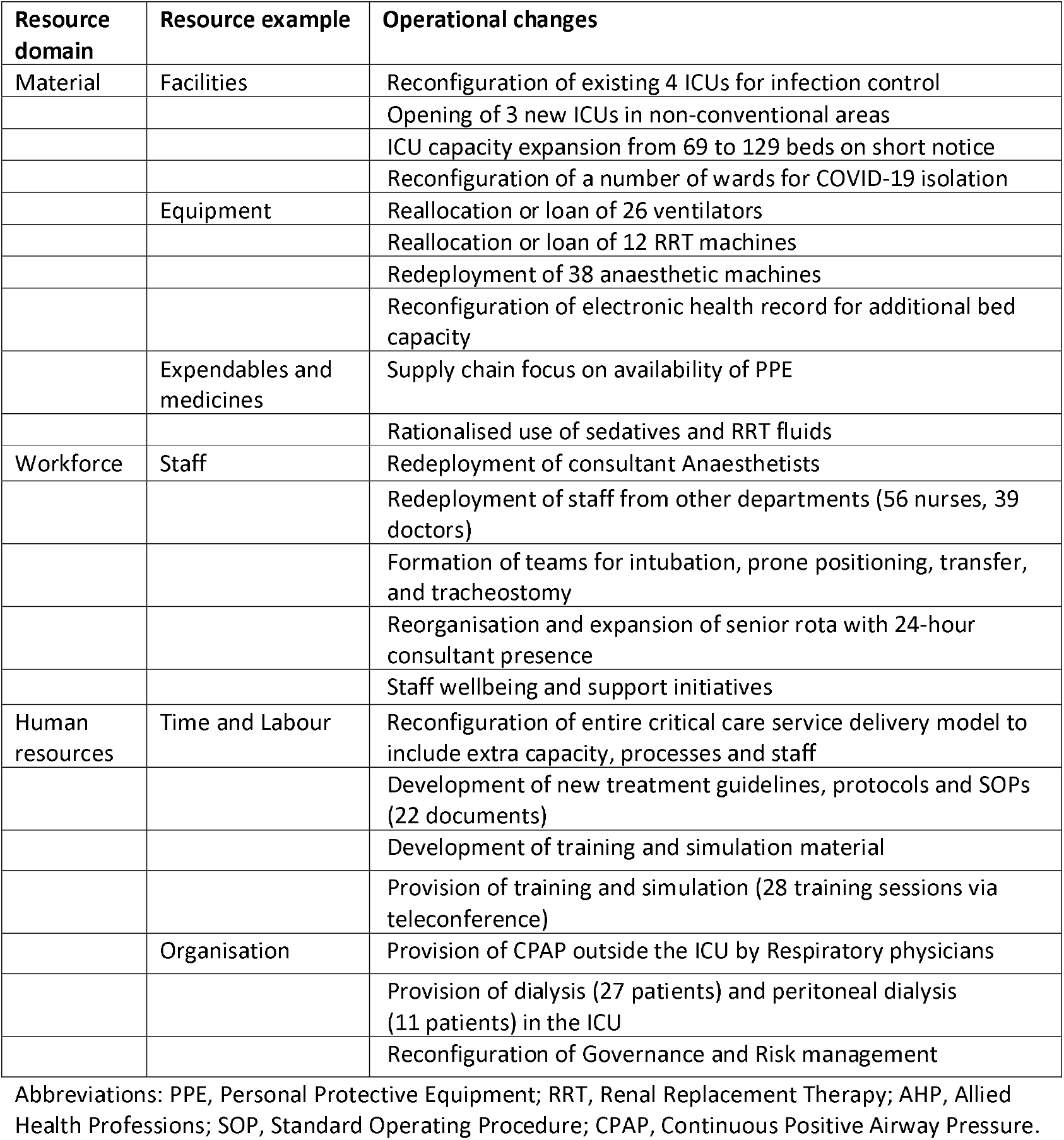
Examples of operational changes across different resource domains over the study period.

| Resource domain | Resource example | Operational changes |
| --- | --- | --- |
| Material | Facilities | Reconfiguration of existing 4 ICUs for infection control |
|  |  | Opening of 3 new ICUs in non-conventional areas |
|  |  | ICU capacity expansion from 69 to 129 beds on short notice |
|  |  | Reconfiguration of a number of wards for COVID-19 isolation |
|  | Equipment | Reallocation or loan of 26 ventilators |
|  |  | Reallocation or loan of 12 RRT machines |
|  |  | Redeployment of 38 anaesthetic machines |
|  |  | Reconfiguration of electronic health record for additional bed capacity |
|  | Expendables and medicines | Supply chain focus on availability of PPE |
|  |  | Rationalised use of sedatives and RRT fluids |
| Workforce | Staff | Redeployment of consultant Anaesthetists |
|  |  | Redeployment of staff from other departments (56 nurses, 39 doctors) |
|  |  | Formation of teams for intubation, prone positioning, transfer, and tracheostomy |
|  |  | Reorganisation and expansion of senior rota with 24-hour consultant presence |
|  |  | Staff wellbeing and support initiatives |
| Human resources | Time and Labour | Reconfiguration of entire critical care service delivery model to include extra capacity, processes and staff |
|  |  | Development of new treatment guidelines, protocols and SOPs (22 documents) |
|  |  | Development of training and simulation material |
|  |  | Provision of training and simulation (28 training sessions via teleconference) |
|  | Organisation | Provision of CPAP outside the ICU by Respiratory physicians |
|  |  | Provision of dialysis (27 patients) and peritoneal dialysis (11 patients) in the ICU |
|  |  | Reconfiguration of Governance and Risk management |

Abbreviations: PPE, Personal Protective Equipment; RRT, Renal Replacement Therapy; AHP, Allied Health Professions; SOP, Standard Operating Procedure; CPAP, Continuous Positive Airway Pressure.

**Figure 3:**
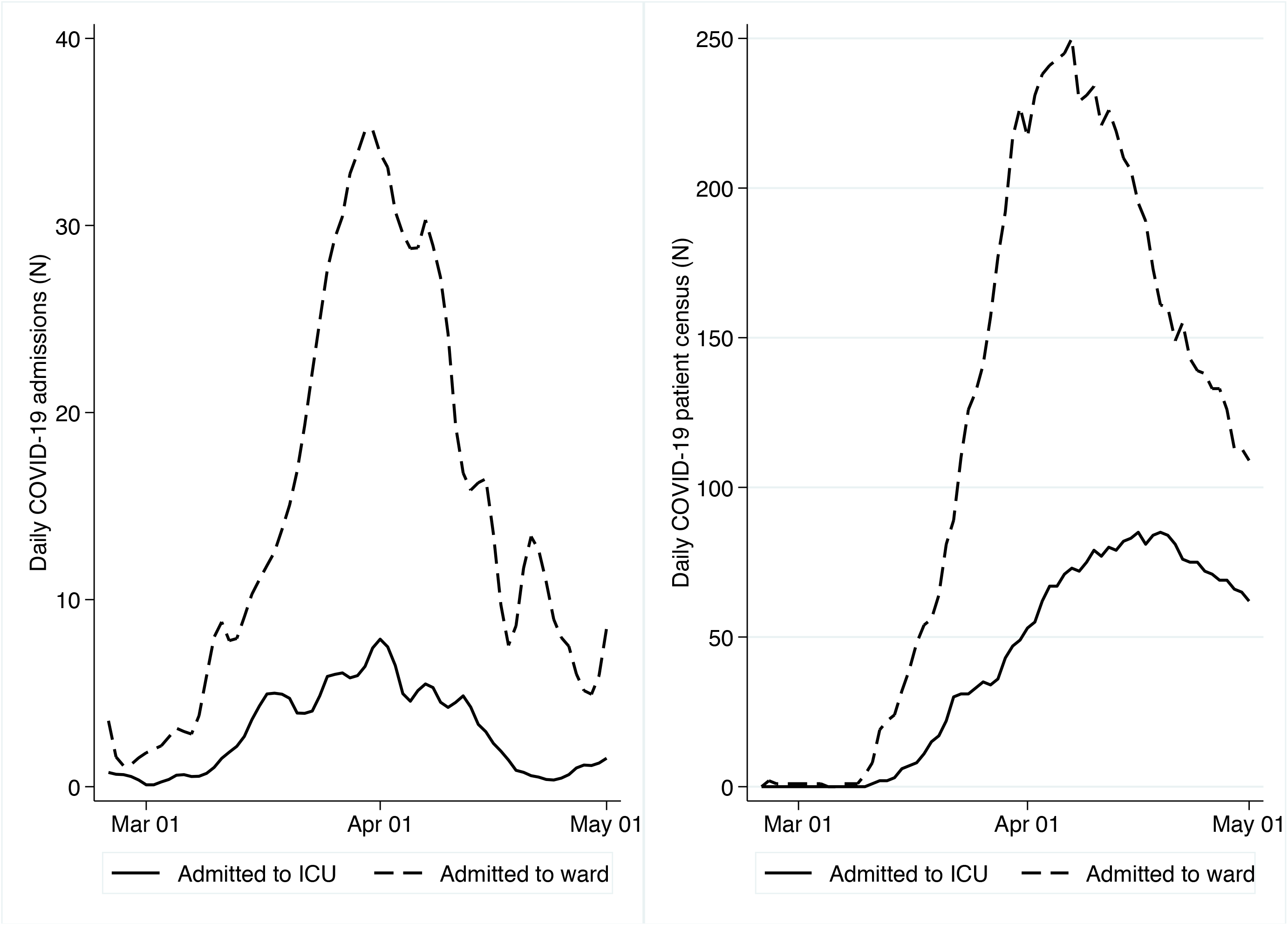
A plot of the surge of patients admitted with COVID-19 over the study period to the ward and the ICU. Left panel: Number of new patients admitted daily. Right panel: Overall number (census) of patients hospitalised with COVID-19.

## DISCUSSION

We describe data from 429 patients hospitalised with COVID-19 in an academic hospital in London UK, of which 76 (18%) were treated in the ICU. We observed a disproportionate burden of COVID-19 hospitalisation and ICU admission in patients from an ethnic minority background, consistent with reports from similarly diverse areas [9] and the rest of the UK [7,8,22]. The range of clinical presentations was broad, similar to reports of Severe Acute Respiratory Syndrome (SARS) and Middle East Respiratory Syndrome (MERS) [23,24]. Respiratory involvement was very frequent, but not universally present. Hence, the reliance on the initial presence of respiratory failure or abnormal chest imaging results to make decisions regarding patient infectiousness and requirement for isolation may be problematic.

Overall hospital mortality was identical to that reported in a recent large UK observational study [7] but patients admitted to the ward had higher morbidity and mortality than previously reported [4,17,25]. A significant proportion experienced clinical deterioration with cardio-respiratory compromise and AKI, meeting the criteria for critical care outreach activation. Patients treated on the ward required considerable input from critical care services, with the ICU outreach team seeing almost four out of ten admitted patients. Similar patients may have been treated in the ICU in other described cohorts and this may explain the difference in unadjusted mortality.

The contribution of the critical care outreach team, as evidenced by the number of patients seen, was crucial in our cohort, however no comparison data exist. As part of their role, outreach teams commonly bridge the gap between ward and ICU by providing expertise, monitoring and interventions outside the ICU environment and, importantly, by engaging in philosophy-of-care discussions with patients and their surrogate decision-makers [26]. In a context of many fixed constraints such as the number of ICU beds, the outreach service is a delivery model that greatly expands the availability of expertise and advanced treatments on the ward. Depending on the institutional context, however, outreach services require appropriate equipment and trained personnel such as doctors, nurses and allied health professionals [26]. This has significant implications on staff planning and resource allocation during a pandemic.

The majority of ward-treated patients in our study had a decision to limit escalation of therapy recorded during their admission, which represents a departure from previous practice. Most patients had treatment limitations placed upon admission. In addition to being clinically frail, serious comorbidities such as active malignancy and stroke were more common in these patients. Within the framework of generic national guidance regarding treatment escalation and ICU admission [27], we believe that the COVID-19 pandemic promoted more proactive communication about goals of care between physicians and their patients. In fact, the Palliative Care service in our institution faced a dramatic increase in ward referrals over the study period [28], but comparative data from other UK settings are lacking. The resource implications of delivering a comprehensive palliative care response during a pandemic are unknown, but likely to be substantial [29-31].

Patients treated in the ICU had high morbidity and mortality. Few of them had trials of CPAP or NIV and almost all required invasive mechanical ventilation, which differs from the experience in the UK [7] and other countries [10]. Pulmonary involvement was significant, with longer periods of mechanical ventilation and more frequent recourse to rescue oxygenation therapies than previously described [9]. Extrapulmonary organ involvement was also common and led to higher utilisation of pharmacological circulatory support and RRT than that reported across the UK [8] and in recent COVID-19 cohorts from Italy (27.8%) [32], the USA (31%) [9], and China (25%) [33]. The high proportion of patients needing organ support in the ICU could be explained by the fact that many patients with less severe organ failure were treated on the ward, with the aid of the outreach service. Similarly, the high incidence of significant extrapulmonary involvement supports the impression that critically ill COVID-19 patients are more similar to those with MERS [24], whereas SARS was a predominantly respiratory disease with single organ failure [34].

Hospital mortality in mechanically ventilated COVID-19 patients was similar to that of unselected cohorts of severe Adult Respiratory Distress Syndrome (ARDS) patients who required ventilation (46.1%), as well as that of mechanically ventilated patients with SARS (45%) and MERS (52.4%) [24,34,35]. In our survival analysis, we identified age, male sex, CKD, elevated CRP, and dyspnoea at presentation as important baseline prognostic factors associated with 30-day mortality after hospitalisation. These are broadly similar to patient characteristics described in reports from other countries [9,36,37] and the UK [7]. We did not, however identify an independent association with ethnicity, which is also in line with findings from a recent large US cohort study [38]. Early identification of patient characteristics associated with poor clinical outcomes and higher resource utilisation is important, when faced with the need to prioritise patients in the face of high demand for critical care services, both in the ICU and on the ward.

The COVID-19 pandemic led to a sudden surge in the overall number of critically ill patients across the hospital, as well as in the intensity and duration of treatments they required. This resulted in significant strain for the ICU in terms of capacity and service delivery. ICU bed capacity was effectively doubled, critically ill patients on the ward required frequent input by the outreach team, while patients in the ICU required prolonged mechanical ventilation and RRT, frequent rescue oxygenation therapies and tracheostomy. To support this increased demand for critical care services, several operational changes were instituted, with support from the Anaesthetic department. These were based on local adaptation of lessons from previous infectious disease outbreaks and information from countries that had been already affected by COVID-19 [2,3,39]. The implementation of this service delivery model required provision of a considerable amount of education and training. Also, these changes were implemented in a short time window and required collaboration across the entire hospital. Our lesson from this process is that clinicians and health administrators need to consider a rapidly scalable model, as health systems could easily become overwhelmed, and our experience is directly comparable to that from other, similarly affected, metropolitan areas [40].

Our study has several strengths. First, it includes a clearly defined cohort with near-complete 30-day outcomes, thus minimising selection bias. Second, it provides a granular clinical description of patient trajectories with previously unknown information regarding ICU outreach involvement and palliative care practice. Third, the inclusion of patients treated both on the ward and in ICU offers a more complete picture of the organ dysfunctions experienced and the corresponding resource implications. Finally, we provide unique details regarding hospital and ICU capacity strain and service reorganisation.

Our study results must, however, be viewed in light of its methodological limitations. The prospective design relied on abstracting data from the EHR and is susceptible to missing data and recording bias. We identified a number of factors associated with mortality but do not imply any causal relationships. Finally, this study was performed in a single centre in the UK and this may limit its generalisability to other institutional contexts.

In conclusion, our study identified age, sex, CKD, baseline CRP and respiratory involvement as factors associated with hospital mortality in COVID-19 patients admitted to a large academic hospital in London, UK. Patients treated on the ward and in the ICU had significant extrapulmonary disease, frequent treatment limitations and a high burden of mortality. Medical wards caring for COVID-19 patients experienced a substantially increased workload and required frequent ICU outreach input. In order to provide effective care under pandemic surge conditions, significant reorganisation took place within the hospital. As a result, when planning an effective response to support patients with COVID-19, clinicians and health administrators should consider the need for additional critical care resources in the ICU, as well as the wider hospital.

## Data Availability

Data sharing is not allowed by the institutional review board.

## Acknowledgements

We thank Professor Alistair McGuire (Department of Health Policy, London School of Economics) for his valuable help in the preparation of the manuscript.

We also thank the King’s College Hospital Business Intelligence Unit for providing us with heatmap images of the pandemic evolution in the area.

We thank “Easy Mobile Forms Software, The DataDyne Group, LLC” for providing 2 months of free access to their data collection tool Magpi.

Finally, the authors would like to thank their patients and fellow health-care workers for providing outstanding patient care at considerable personal risk.

## Author contributions

SV and RM conceived the study and its design, had full access to the data, and take responsibility for the integrity of the data and accuracy of the analysis. SV, AW, VM, SC, CLS, JP, KL, TP, CS, KA, BA, RB, FJ, SAH, WB and RM extracted and entered data. SV and RM contributed to data analyses. SV, AW, VM, SC, CLS, JP, KL, WB and RM contributed to data interpretation. SV and RM drafted the manuscript. All authors critically revised the drafted manuscript and approve of the submitted manuscript.

## REFERENCES

1. World Health Organization. Situation Report-133 June 1, 2020. https://www.who.int/docs/default-source/coronaviruse/situation-reports/20200601-covid-19-sitrep-133.pdf?sfvrsn=9a56f2ac_4 (accessed July 7, 2020).

2. Wu Z, McGoogan JM. Characteristics of and Important Lessons From the Coronavirus Disease 2019 (COVID-19) Outbreak in China: Summary of a Report of 72314 Cases From the Chinese Center for Disease Control and Prevention. JAMA. 2020 (In Press). https://doi.org/10.1001/jama.2020.2648.

3. Grasselli G, Pesenti A, Cecconi M. Critical Care Utilization for the COVID-19 Outbreak in Lombardy, Italy: Early Experience and Forecast During an Emergency Response. JAMA 2020; 323: 1545–6.

4. Richardson S, Hirsch JS, Narasimhan M, et al. Presenting Characteristics, Comorbidities, and Outcomes Among 5700 Patients Hospitalized With COVID-19 in the New York City Area. JAMA 2020; 323: 2052–9.

5. Public Health England. Daily UK COVID-19 Cases Update, 2020. https://coronavirus.data.gov.uk (accessed July 7, 2020).

6. Armstrong RA, Kane AD, Cook TM. Outcomes from intensive care in patients with COVID-19: a systematic review and meta-analysis of observational studies. Anaesthesia 2020 (In Press). https://doi.org/10.1111/anae.15201.

7. Docherty AB, Harrison EM, Green CA, et al. Features of 20 133 UK patients in hospital with covid-19 using the ISARIC WHO Clinical Characterisation Protocol: prospective observational cohort study. BMJ 2020; 369: m1985.

8. Intensive Care National Audit&Research Centre. ICNARC report on COVID-19 in critical care, 2020. https://www.icnarc.org/Our-Audit/Audits/Cmp/Reports (accessed July 1, 2020).

9. Cummings MJ, Baldwin MR, Abrams D, et al. Epidemiology, clinical course, and outcomes of critically ill adults with COVID-19 in New York City: a prospective cohort study. Lancet 2020; 395: 1763–70.

10. Grasselli G, Zangrillo A, Zanella A, et al. Baseline Characteristics and Outcomes of 1591 Patients Infected With SARS-CoV-2 Admitted to ICUs of the Lombardy Region, Italy. JAMA 2020; 323: 1574–81.

11. World Health Organization. Clinical management of severe acute respiratory infection when COVID-19 is suspected (v1.2). https://www.who.int/publications-detail/clinical-management-of-severe-acute-respiratory-infection-when-novel-coronavirus-(ncov)-infection-is-suspected (accessed April 20, 2020).

12. von Elm E, Altman DG, Egger M, et al. The Strengthening the Reporting of Observational Studies in Epidemiology (STROBE) Statement: guidelines for reporting observational studies. Int J Surg 2014; 12: 1495–9.

13. Magpi. Easy Mobile Forms Software. Washington DC: The DataDyne Group, LLC, 2020.

14. Ministry of Housing Communities & Local Government. English indices of deprivation 2019. https://www.gov.uk/government/statistics/english-indices-of-deprivation-2019 (accessed May 15, 2020).

15. Smith GB, Redfern OC, Pimentel MA, et al. The National Early Warning Score 2 (NEWS2). Clinical medicine (London, England) 2019; 19: 260.

16. Vincent JL, Moreno R, Takala J, et al. The SOFA (Sepsis-related Organ Failure Assessment) score to describe organ dysfunction/failure. On behalf of the Working Group on Sepsis-Related Problems of the European Society of Intensive Care Medicine. Intensive Care Med 1996; 22: 707–10.

17. Myers LC, Parodi SM, Escobar GJ, Liu VX. Characteristics of Hospitalized Adults With COVID-19 in an Integrated Health Care System in California. JAMA 2020; 323: 2195–8.

18. Rockwood K, Song X, MacKnight C, et al. A global clinical measure of fitness and frailty in elderly people. CMAJ 2005; 173: 489–95.

19. Muscedere J, Waters B, Varambally A, et al. The impact of frailty on intensive care unit outcomes: a systematic review and meta-analysis. Intensive Care Med 2017; 43: 1105–22.

20. Obolensky L, Clark T, Matthew G, Mercer M. A patient and relative centred evaluation of treatment escalation plans: a replacement for the do-not-resuscitate process. J Med Ethics 2010; 36: 518–20.

21. KDIGO. Section 2: AKI Definition. Kidney international supplements 2012; 2: 19–36.

22. Lassale C, Gaye B, Hamer M, Gale CR, David Batty G. Ethnic Disparities in Hospitalisation for COVID-19 in England: The Role of Socioeconomic Factors, Mental Health, and Inflammatory and Pro-inflammatory Factors in a Community-based Cohort Study. Brain Behav Immun. 2020 (In Press). https://doi.org/10.1016/j.bbi.2020.05.074.

23. Peiris JS, Yuen KY, Osterhaus AD, Stohr K. The severe acute respiratory syndrome. N Engl J Med 2003; 349: 2431–41.

24. Arabi YM, Al-Omari A, Mandourah Y, et al. Critically Ill Patients With the Middle East Respiratory Syndrome: A Multicenter Retrospective Cohort Study. Crit Care Med 2017; 45: 1683–95.

25. Guan WJ, Ni ZY, Hu Y, et al. Clinical Characteristics of Coronavirus Disease 2019 in China. N Engl J Med 2020; 382: 1708–20.

26. Jones DA, DeVita MA, Bellomo R. Rapid-response teams. N Engl J Med 2011; 365: 139–46.

27. Royal College of Physicians. Ethical dimensions of COVID-19 for frontline staff, 2020. https://www.rcplondon.ac.uk/news/ethical-guidance-published-frontline-staff-dealing-pandemic (accessed April 18, 2020).

28. Lovell N, Maddocks M, Etkind SN, et al. Characteristics, Symptom Management, and Outcomes of 101 Patients With COVID-19 Referred for Hospital Palliative Care. Journal of Pain and Symptom Management 2020; 60: e77–e81.

29. Sprung CL, Joynt GM, Christian MD, Truog RD, Rello J, Nates JL. Adult ICU Triage During the Coronavirus Disease 2019 Pandemic: Who Will Live and Who Will Die? Recommendations to Improve Survival. Critical Care Medicine. 2020 (In Press). https://journals.lww.com/ccmjournal/Fulltext/9000/Adult_ICU_Triage_During_the_Coronavirus_Disease.95654.aspx.

30. May P, Normand C, Cassel JB, et al. Economics of Palliative Care for Hospitalized Adults With Serious Illness: A Meta-analysis. JAMA Intern Med 2018; 178: 820–9.

31. Fausto J, Hirano L, Lam D, et al. Creating a Palliative Care Inpatient Response Plan for COVID-19-The UW Medicine Experience. J Pain Symptom Manage 2020: S0885-3924(20)30176-7.

32. Fanelli V, Fiorentino M, Cantaluppi V, et al. Acute kidney injury in SARS-CoV-2 infected patients. Crit Care 2020; 24: 155.

33. Yang X, Yu Y, Xu J, et al. Clinical course and outcomes of critically ill patients with SARS-CoV-2 pneumonia in Wuhan, China: a single-centered, retrospective, observational study. Lancet Respir Med 2020; 8: 475–81.

34. Fowler RA, Lapinsky SE, Hallett D, et al. Critically ill patients with severe acute respiratory syndrome. JAMA 2003; 290: 367–73.

35. Bellani G, Laffey JG, Pham T, et al. Epidemiology, Patterns of Care, and Mortality for Patients With Acute Respiratory Distress Syndrome in Intensive Care Units in 50 Countries. JAMA 2016; 315: 788–800.

36. Liang W, Liang H, Ou L, et al. Development and Validation of a Clinical Risk Score to Predict the Occurrence of Critical Illness in Hospitalized Patients With COVID-19. JAMA Intern Med. 2020 (In Press). https://doi.org/10.1001/jamainternmed.2020.2033.

37. Wu C, Chen X, Cai Y, et al. Risk Factors Associated With Acute Respiratory Distress Syndrome and Death in Patients With Coronavirus Disease 2019 Pneumonia in Wuhan, China. JAMA Intern Med. 2020 (In Press). https://doi.org/10.1001/jamainternmed.2020.0994.

38. Price-Haywood EG, Burton J, Fort D, Seoane L. Hospitalization and Mortality among Black Patients and White Patients with Covid-19. N Engl J Med 2020; 382: 2534–43.

39. Kain T, Fowler R. Preparing intensive care for the next pandemic influenza. Crit Care 2019; 23: 337.

40. Griffin KM, Karas MG, Ivascu NS, Lief L. Hospital Preparedness for COVID-19: A Practical Guide from a Critical Care Perspective. Am J Respir Crit Care Med 2020; 201: 1337–44.

